# Transcriptional profiling of antidepressant ketamine and electroconvulsive therapy treatment

**DOI:** 10.1101/2025.07.29.25332162

**Authors:** Artemis Zavaliangos-Petropulu, Ginny Ghang, Toni Boltz, Paloma Pfeiffer, Lingyu Zhan, Eliza Congdon, Randall T Espinoza, Katherine L Narr, Roel A Ophoff

**Affiliations:** Ahmanson-Lovelace Brain Mapping Center, Department of Neurology, Geffen School of Medicine at the University of California, Los Angeles, CA, USA; Department of Human Genetics, David Geffen School of Medicine, University of California Los Angeles, Los Angeles, CA, USA; Jane and Terry Semel Institute for Neuroscience and Human Behavior, Department of Psychiatry and Biobehavioral Sciences, Geffen School of Medicine at the University of California, Los Angeles, CA, USA

**Keywords:** RNA-seq, gene expression, depression, ketamine, electroconvulsive therapy

## Abstract

1.

**Background:** Treatment-resistant depression (TRD) affects 30-50% of patients with major depressive disorder (MDD). Electroconvulsive therapy (ECT) and sub-anesthetic ketamine treatment can relieve TRD, yet their antidepressant mechanisms remain unclear. Peripheral blood gene expression offers a non-invasive proxy to examine potential treatment-response biomarkers.

**Methods:** We conducted a transcriptome analysis on peripheral blood samples from individuals with TRD undergoing ECT (N=37) or serial ketamine infusions (N=60), and non-depressed controls (N=35). Samples were collected at baseline and at multiple follow-up time points. Differential gene expression (DGE) at the single gene and network level identified transcriptional changes and co-regulated gene modules associated with diagnosis, treatment, and remission status using Weighted Gene Co-Expression Network Analysis (WGCNA), including correction for multiple comparisons.

**Results:** Longitudinal transcriptional changes were not detected for either treatment for individual genes or networks (FDR corrected or |logFC|>0.05). When comparing remitters and non-remitters at baseline in the ketamine group, we observed evidence of enrichment for immune-related functions overall with one gene significantly differentially expressed (i.e., IGKV1-9) (p=2.5E-05, logFC=-0.51). In the ECT sample, when considering gene networks, we observed significant interaction effects between time and diagnosis. At least six co-regulated gene modules yielded significant differences at baseline between ECT patients and controls.

**Conclusion:** Despite the robust clinical improvements associated with ECT and ketamine, peripheral blood RNA-seq revealed limited detectable longitudinal gene expression changes. However, pre-treatment differences in gene expression profiles suggest some potential predictive value. Larger samples are warranted to clarify peripheral molecular signatures of rapid-acting antidepressant response.

## 2. Introduction

Major depressive disorder (MDD) is a debilitating psychiatric disorder, affecting over 250 million people worldwide(1). Despite advancements in pharmacological and psychotherapeutic interventions, a significant proportion of patients—between 30-50%—do not benefit from conventional treatments, a condition known as treatment-resistant depression (TRD)(2,3). TRD poses considerable challenges(4), including increased risks of chronic illness(5), disability(6), and suicidal behavior(7), necessitating alternative therapeutic strategies.

Electroconvulsive therapy (ECT)(8–10) and ketamine(11,12)(13) are both rapid-acting alternative treatment options for patients with TRD. ECT, which induces a brief seizure through electrical stimulation of the brain, has long been considered amongst the most effective interventions for severe depression(14). Ketamine, traditionally used as an anesthetic, has since been recognized for its rapid and pronounced antidepressant effects when administered in subanesthetic doses(15). Despite progress(16,17), the molecular mechanisms underlying ECT and ketamine’s antidepressant effects remain largely unknown.

Preclinical brain-derived gene expression studies have provided important insights into the molecular mechanisms potentially underlying the antidepressant effects of ECT(18–20) and ketamine(21–23). Unfortunately, experiments requiring access to brain tissue in living humans are not feasible. However, although the brain and peripheral blood exhibit distinct transcriptional profiles(24), prior data shows a meaningful overlap in the expression of genes implicated in depression, suggesting that more accessible sampling of peripheral blood may also reveal CNS mechanisms(25,26). For instance, findings suggest that 35%-80% of known transcripts are detectable in both brain and blood tissue samples(25), supporting the utility of using peripheral blood as a proxy for some aspects of brain gene expression. Several depression-related genes and pathways, particularly those involved in inflammation and immune regulation, are similarly dysregulated in brain tissue and peripheral blood(27). This overlap provides further rationale for using peripheral blood to investigate gene expression changes that, while potentially distinct in the brain, may still reflect biomarkers and treatment response mechanisms in psychiatric disorders.

Since ECT and ketamine both exhibit robust behavioral effects, these treatment modalities present an opportunity to evaluate peripheral blood gene expression molecular signatures linked with outcomes and brain-derived phenotypes. Very few prior studies have explored peripheral blood gene expression in humans within the context of fast-acting antidepressants(28–30). Here, we performed a whole-blood transcriptome analysis (using RNA sequencing) of patients with TRD undergoing either naturalistic, open-label ECT or ketamine treatment. The overall goal was to identify differential gene expression (DGE) genes and molecular pathways associated with rapidly acting antidepressant response for either treatment modality. By exploring the transcriptional landscape in peripheral blood, we expected to identify gene changes that would elucidate the mechanisms of rapid therapeutic response and identify blood-based biomarkers that could potentially inform treatment outcomes, thereby contributing to the ongoing efforts to personalize interventions for depression.

## 3. Methods

Data for this study were collected under a NIMH-supported Connectomes Related to Human Disease (CRHD) U01 grant (MH110008)(31). The primary aim of the project was to identify connectome-based imaging correlates and predictors of treatment response to three rapid-acting antidepressant interventions: ECT, serial ketamine, and total sleep deprivation. Peripheral blood for RNA-sequencing was collected as a secondary objective, using two PaxGene RNA tubes per blood draw at set time points. Tubes were incubated at room temperature for 2 hours, then frozen and transported to the UCLA Pathology Research Portal for long-term storage, pending batch extraction and quality control by the UCLA Neuroscience Genomics Core.

### 3.1 Participants

Patients receiving ketamine and healthy control participants were recruited from the Los Angeles area through advertisements or clinician referral, or clinicaltrials.gov (NCT02165449) for ketamine specifically. ECT participants were recruited from individuals already scheduled to receive ECT as part of routine care at the Resnick Neuropsychiatric Hospital at UCLA. All patient and control participants received diagnostic evaluation using the Structured Clinical Interview for DSM-5. Inclusion criteria for all subjects recruited were age between 20-46 years, able to read and write English and provide informed consent. Exclusion criteria for all participants were dementia, schizophrenia spectrum disorders, or a psychotic disorder due to a general medical condition, neurological disorder, or other serious medical condition, current or history of substance abuse (within the last three months) and pregnancy, each confirmed by blood and urine laboratory tests. TRD patients receiving Ketamine or ECT were recruited independently (not randomized), though their inclusion/exclusion criteria were similar as described below.

#### 3.1.1 Ketamine participants

Participants receiving ketamine met DSM-5 criteria for a major depressive episode (MDE) (unipolar or bipolar depression with/without psychotic features) and TRD, defined as not having achieved adequate response following ≥2 antidepressant trials of sufficient dosage for > 4-6 weeks each and having experienced continuous depression for >6 months. Inclusion required moderate to severe baseline depressive symptoms (17-Hamilton Depression Rating Scale (HDRS) score of 17 or higher), and stable use of antidepressants or mood stabilizers for >6 weeks before enrollment. Additional exclusion criteria included history of seizures or withdrawal-related convulsions, prior neuromodulation treatment or ketamine within the last six months, past psychotic reactions to medications and any other contraindications to ketamine. Patients were also required to taper from any use of benzodiazepines (as judged by psychiatrist) >24 hours prior to treatment sessions.

#### 3.1.2 ECT participants

Eligibility for ECT itself was independently verified based on clinical indications according to the APA ECT Task Force Report guidelines. ECT treatment protocols were not manipulated and was provided as clinically indicated. Inclusion and exclusion criteria were otherwise the same as for the ketamine group. However, ECT patients discontinued medications, including antidepressants and anticonvulsants, for at least 48 to 72 hours prior to initiation of ECT and study enrollment.

#### 3.1.3 Controls

Unaffected controls did not receive any treatment. A subset of control subjects had data collected at a second time point within a similar time frame as the post-fourth infusion ketamine time point (2 weeks). Additional exclusion criteria for controls were any current or past psychiatric disorders or use of any antidepressants or mood stabilizers within the past 6 months. All participants gave written informed consent in accordance with procedures approved by the UCLA Institutional Review Board (IRB).

### 3.2 Study Design

#### 3.2.1 Ketamine treatment

Using an open-label design, patients received an intravenous infusion of racemic ketamine (0.5mg/kg) administered over 40 minutes, four times over two weeks. Blood was collected at baseline, 24 hours following the first infusion, 24 hours following the fourth infusion, and at a 5 – week follow-up (**Figure 1**). All controls had blood collected at baseline and a subset of controls had blood collected again two weeks later.

**Figure 1.**
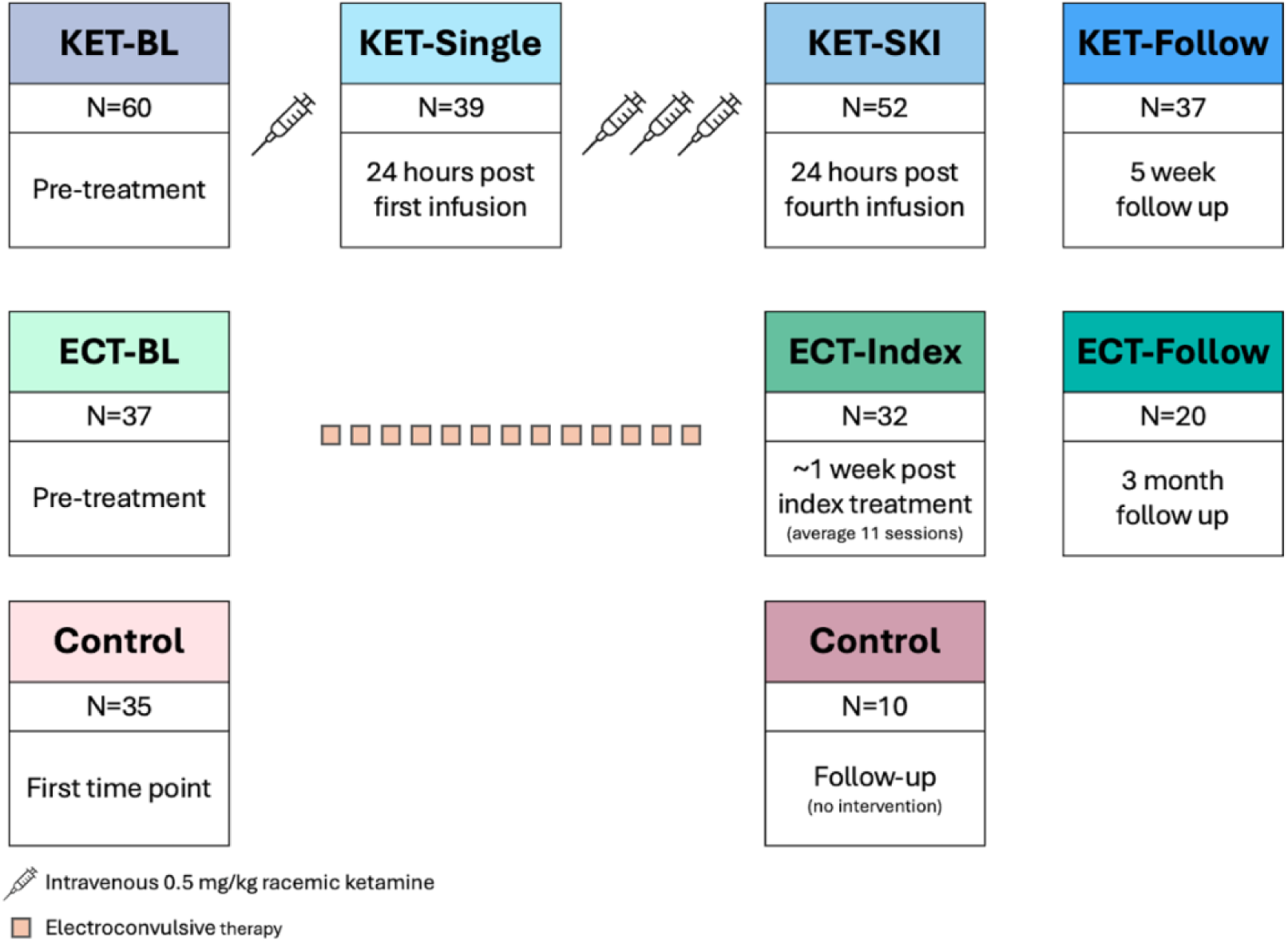
Study design for ketamine (KET) and ECT blood data collection, including sample sizes and description of time points. Demographics for full sample can be found in **Table 1**, and different sample sizes across time points can be found in **Supplemental Table 1**.

**Table 1.** Demographics for the ketamine and ECT samples and healthy controls (HC), with each treatment group compared with HCs separately. Additional information about race, education, concurrent medications, and other psychiatric comorbidities can be found in **Supplemental** Tables 2-3.

|  | TRD | HC | P-value |
| --- | --- | --- | --- |
| *Ketamine* |  |  |  |
| N | 60 | 35 | - |
| Age | 39.5±11.2 | 33.5±12 | 0.019 |
| Sex (Percent female) | 46.7% | 54.3% | 0.6 |
| Baseline HDRS | 18.8±4.75 | - | - |
| Duration of Illness (years) | 3.9±5.9 | - | - |
| *ECT* |  |  |  |
| N | 37 | 35 | - |
| Age | 41.6±16.0 | 33.5±12 | 0.018 |
| Sex (Percent female) | 67.6% | 54.3% | 0.36 |
| Baseline HDRS | 21.3±6.7 | - | - |
| Duration of Illness (years) | 2.9±6.9 | - | - |

#### 3.2.2 ECT treatment

Using a naturalistic design, eligible patients received a clinically prescribed course of ECT (5000Q MECTA Corp., Tualatin, Oregon), administered three times per week. Anesthesia and muscle relaxation were achieved using methohexital (1 mg/kg) and succinylcholine (1 mg/kg). ECT followed the seizure threshold (ST) titration method. Right-unilateral ECT was delivered at five times the ST with an ultrabrief pulse width (0.3 ms), while bilateral ECT was given at 1.5 times ST with a brief pulse width (0.5 ms). All patients initially received RUL ECT, but ∼30% transitioned to bilateral lead placement based on clinical judgement of response rates. On average, patients underwent 10.9±3.9 ECT sessions, with an average ST of 29.57±25.94 mC and an average seizure duration of 67.8±19.5 seconds across all ECT treatments. Patients were evaluated at three time points: within 24 hours before the first ECT session (baseline), within one week of completing the ECT index series occurring ∼3-4 weeks after treatment initiation, and three months after ECT treatment ended or symptom relapse, whichever occurred first (**Figure 1**).

### 3.3 RNA-Sequencing

RNA sequencing was performed by the UCLA Neuroscience Genomics Core using an Illumina sequencer, with library preparation conducted using the TruSeq + RiboZero Gold protocol. RNA-seq data quality was visually assessed using FastQC(32), which provided comprehensive quality control metrics for each sample, including per-base sequence quality, GC content, and adapter contamination. Samples failing initial quality checks were excluded from the analysis. Kallisto(33) was used to pseudoalign reads to the GRCh38 gencode transcriptome and quantify estimates of transcript expression. Samples with RNA Integrity Number (RIN) <6 were excluded. Genetic data for this study can be found in the Gene Expression Omnibus database repository.

### 3.4 Data Analysis

Kallisto outputs were read into R version 4.3.0 gcc 10.2.0 and analyzed as transcripts per million (TPM). We filtered out non–protein-coding genes and genes <10 reads per million mapped in at least two libraries, resulting in a different number of genes analyzed for each dataset due to varying sample sizes (**Table 2**). TPM values were (log2+1)-transformed and analyzed using *limma-voom*(34). *DuplicateCorrelation* was used to adjust for within-subject variability when performing longitudinal analyses. The analysis yielded similar results using DESeq2(35). R-package *WGCNA*(36) was used to perform Weighted Gene Co-Expression Network Analysis (WGCNA) to identify gene co-expression networks with a minimum module size of 30. Multidimensional scaling (MDS) plots, mean-variance j-plots, and volcano plots can be found in **Supplemental Material**.

**Table 2.**
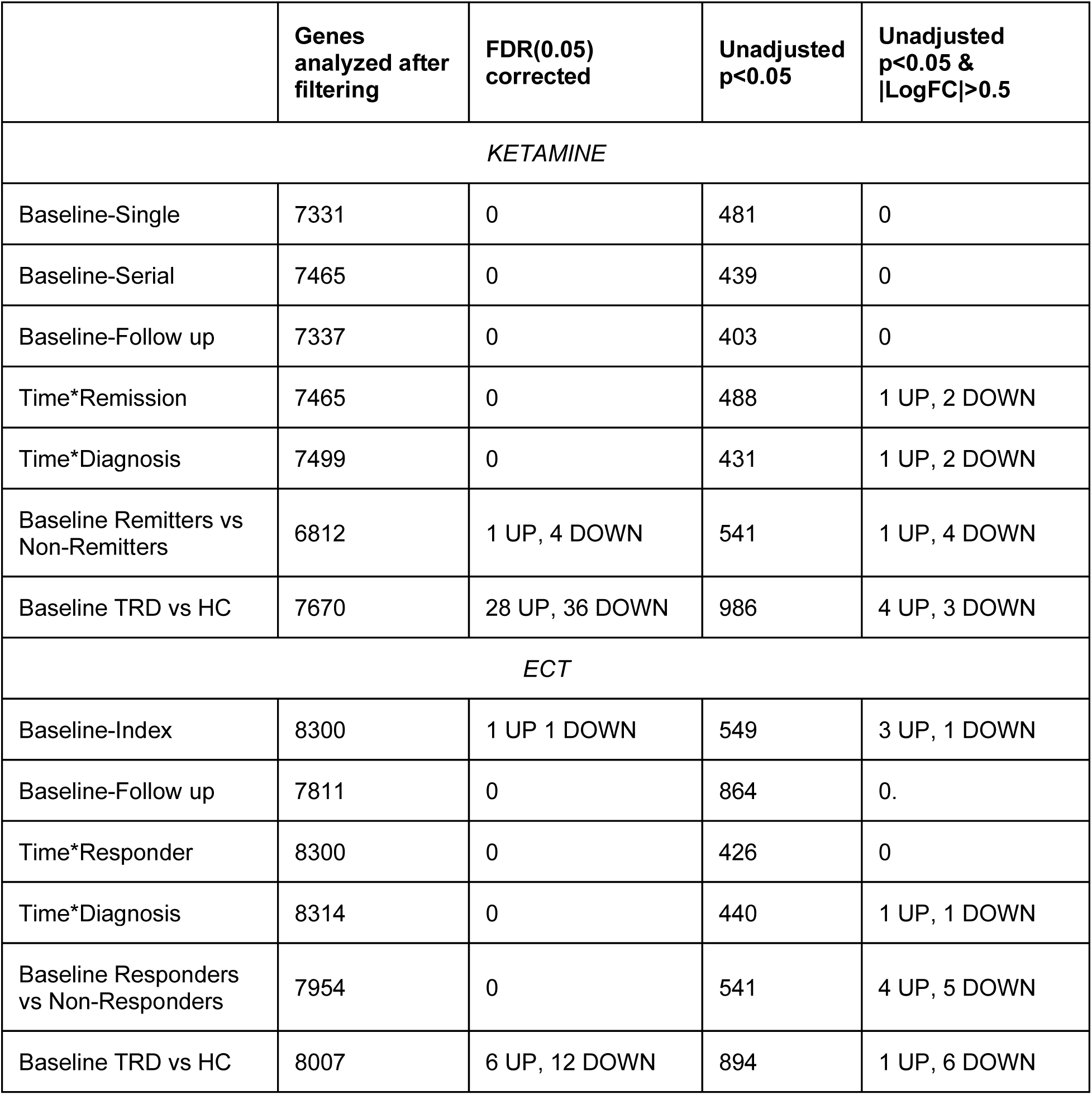
Summary of DGE findings. The number of genes analyzed and their significance thresholds differ for pairwise comparisons between baseline-single infusion, baseline-serial infusion, and remitters vs non-remitters at baseline.

#### 3.4.1 Differential Gene Expression (DGE)

##### 3.4.1.1 Ketamine DGE

We performed pairwise comparisons across time points in patients receiving ketamine treatment, adjusting for age, sex, RIN and the first 10 principal components (PC). Pairwise comparisons included 1) baseline to post-first (single) infusion, 2) baseline to post-fourth infusion and 3) baseline to follow-up. To evaluate effects of remission status, remitters were classified as patients achieving a post-fourth infusion HDRS score of ≤7. A mixed-effect model tested for time*remission interaction by comparing baseline to post-fourth infusion. We similarly tested for a time*diagnosis interaction including controls with longitudinal data. We also performed a DGE analysis comparing baseline gene expression in remitters compared to non-remitters, and patients compared to controls. All analyses were corrected for multiple comparisons using False Discovery Rate (FDR(0.05))(37). Functional classification of findings were determined using Gene Ontology for results with an absolute value of log fold change (logFC)(38) greater than 0.5 and uncorrected p-values<0.05.

##### 3.4.1.2 ECT DGE

Similar to the ketamine analysis, we performed pairwise comparisons across time points in patients receiving ECT, adjusting for age, sex, RIN and the first 10 PCs. ECT pairwise comparisons included 1) baseline to post-index and 2) baseline to follow-up. Clinical response was defined as a ≥30% reduction in HDRS scores post-index, following the widely used criterion for a meaningful response(23). Time*remission and time*diagnosis models included baseline and post-index time points.

#### 3.4.2 Network Analysis

WGCNA modules were generated using all data points with RIN≥6 from the controls, ketamine, and ECT samples. Eigengenes for each module were calculated for individual samples. In the ketamine sample, limma was used to test for associations between eigengenes and time, time*remission interaction, time*diagnosis, and remission status and diagnosis at baseline. Likewise, in the ECT sample, limma was used to test for associations between eigengenes and time, time*response, and time*diagnosis interactions, and response status and diagnosis at baseline.

## 4. Results

Sixty-five participants were enrolled for ketamine infusions, but after filtering for RIN, N=60 were analyzed. By the fourth infusion, 48% of participants receiving ketamine achieved remission (HDRS≤7). Forty-one participants were enrolled for ECT, but after filtering, N=37 were analyzed. Post-Index ECT, 49% of participants were classified as responders (≥30% decrease in HDRS from baseline to post-index). **Table 1**. provides demographic and clinical information for each sample at baseline.

### 4.1 DGE Analysis

#### 4.1.1 Ketamine DGE analysis

No significant transcriptional changes were detected in the ketamine group after multiple comparison correction (**Table 2**). Trending (uncorrected p<0.05, |logFC|>0.5) interactions for time*remission status were observed in 3 genes, and an additional 3 genes for time*diagnosis when compared to controls. Since none survived FDR, gene ontology analysis was not performed. Baseline FDR-corrected analysis identified 1 upregulated and 4 downregulated genes in remitters vs. non-remitters. However, only one of these genes (IGKV1-9) had a significant logFC (p=0.000025, logFC=-0.51). Functional classification suggests this gene is involved in molecular functions related to immune response and immunoglobulin production. When comparing baseline gene expression of all patients receiving ketamine to controls, 64 genes were differentially expressed after FDR (**Figure 2**). However when applying the logFC threshold, only 1 upregulated gene (RPL36AP37, adjusted p=0.02, logFC=0.53) and one down regulated gene (RNF144B, adjusted p=0.016, logFC=-0.62) remained; functional annotation revealed that RPL36AP37 is a ribosomal pseudogene with no known protein-coding function, while RNF144B encodes an E3 ubiquitin ligase implicated in the regulation of apoptosis and inflammatory signaling pathways.

**Figure 2.**
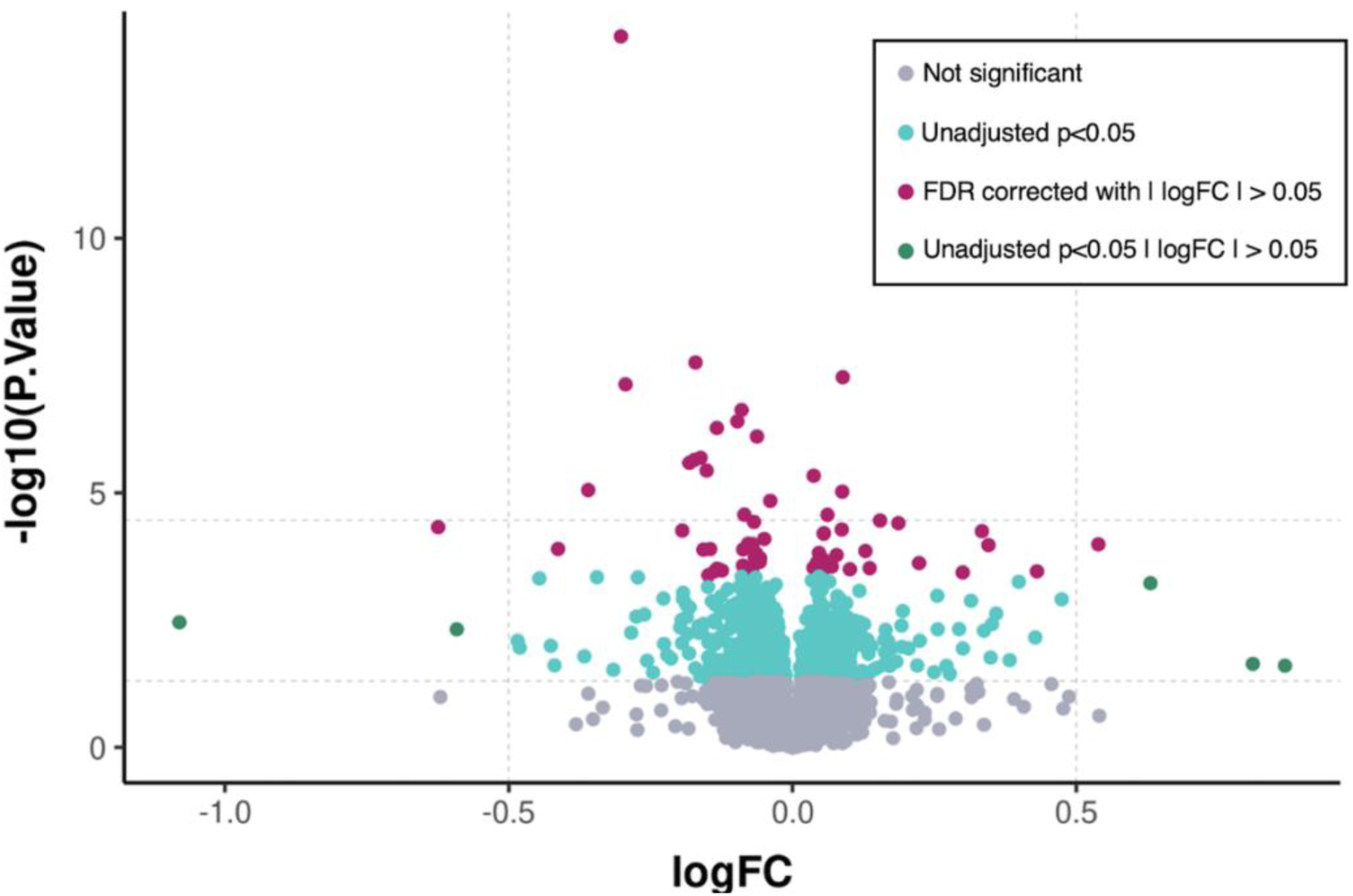
Volcano plot for baseline differential gene expression comparing HC and patients receiving ketamine.

#### 4.1.2 ECT DGE analysis

Two genes showed significant change after FDR correction following ECT when comparing baseline to post-index treatment. RPL26P5 was downregulated post-index when compared to baseline (FDR adjusted p=0.008, logFC=-0.15) and EIF1AY was upregulated post-index compared to baseline (FDR-adjusted p=0.008, logFC=0.23), though neither had a logFC greater than 0.5 or smaller than –0.5. After FDR correction, no significant interactions were observed between the change to post-index with treatment response or diagnosis when compared to controls. Significant changes were also absent from baseline to follow-up. At baseline, no significant DGE was observed between responders and non-responders after FDR correction. When comparing pre-treatment patients receiving ECT and controls, 18 genes were differentially expressed (**Figure 3)**, however, only 1 gene had a substantial logFC (HLA-DQA1, adjusted p=0.02, logFC=-0.99); gene ontology analysis identified its role in antigen processing and presentation via MHC class II.

**Figure 3.**
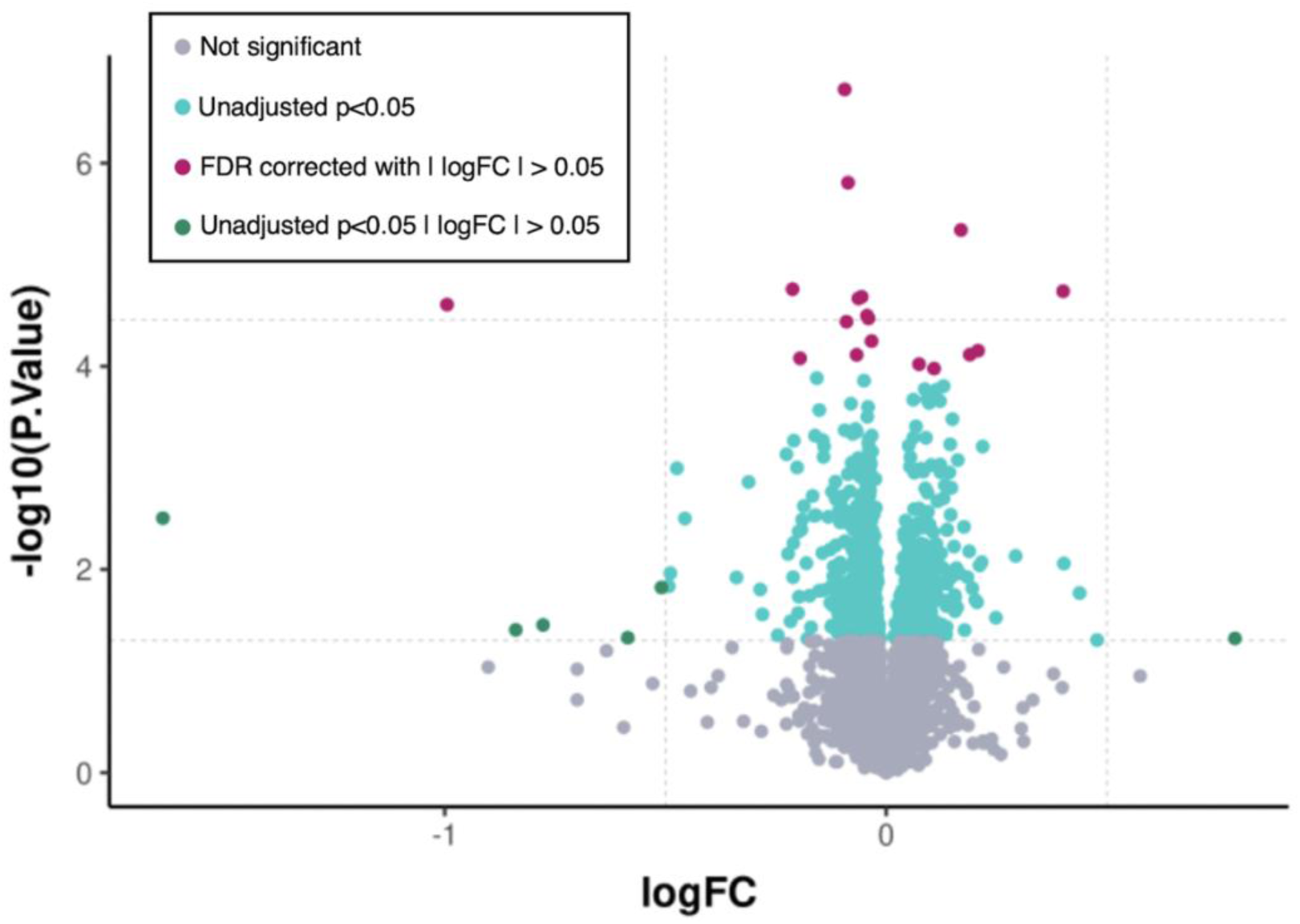
Volcano plot for baseline differential gene expression comparing HC and patients receiving ECT.

### 4.2 Network Analysis

#### 4.2.1 Ketamine network analysis

Network analysis revealed no significant associations with module eigengenes (ME) for any analyses after FDR correction. Nevertheless, trending associations (unadjusted p<0.05) were observed in ME21 for a time*remission interaction, where remitters showed a modest increase over time while non-remitters remained stable. For baseline comparisons, four modules showed trending differences by remission status (ME21, ME15, ME12, and ME10) and one module by diagnosis (ME22) **Figure 4**. In an exploratory analysis, we performed gene ontology in genes included in ME21 and found enrichment for genes involved in molecular functions related to immune response and immunoglobulin production, primarily driven by IGKV1-9, which was the gene identified in DGE analysis. Full gene lists for trending modules and gene ontology can be found in **Supplemental Material.**

**Figure 4.**
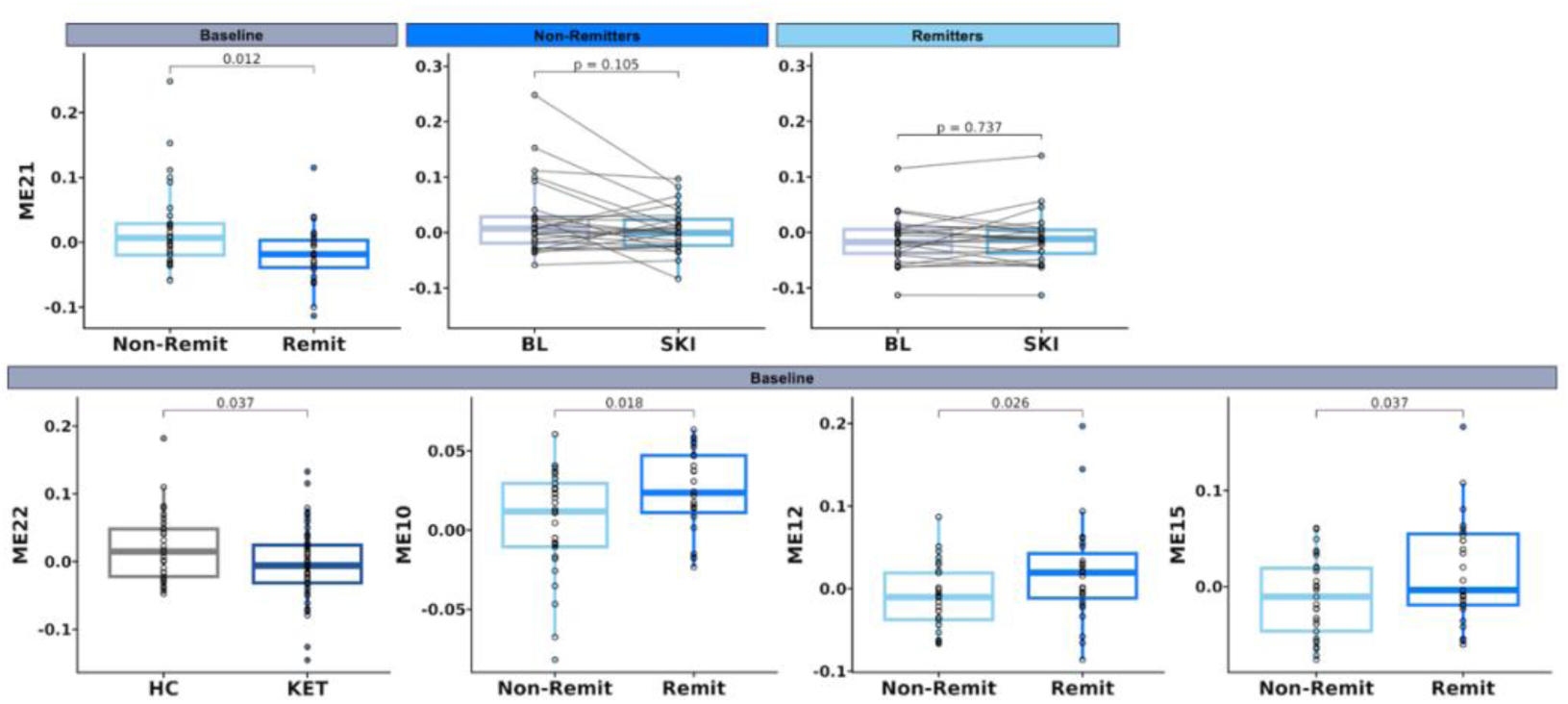
Trending eigengene results within the ketamine sample. A time-by-remission interaction was observed for ME21 (*p*<0.05), with remitters showing a modest post-treatment increase. At baseline, ME21, ME15, ME12, and ME10 differed by remission status, and ME22 differed by diagnosis. All *p*-values are unadjusted.

#### 4.2.2 ECT network analysis

A significant time*diagnosis interaction was observed in 8 modules (ME3, ME1, ME14, ME2, ME10, ME11, ME21, ME16) after correcting for FDR (**Table 3**), however post-hoc analyses revealed no notable significant changes in ECT or controls (**Supplemental Figure 13**). At baseline, 12 eigengenes significantly differed between ECT patients and controls (ME14, ME2, ME23, ME10, ME13, ME3, ME15, ME9, ME7, ME21, ME11, ME1; **Table 3, Supplemental Figure 15**). ME11 differed when comparing baseline responders and non responders but did not pass FDR correction.

**Table 3.** WGCNA network analysis findings.

| Module | Model | logFC | Raw p-value | Adjusted p-value |
| --- | --- | --- | --- | --- |
| *Ketamine* |  |  |  |  |
| ME21 | time*remission | -0.04 | 0.03 | 0.7 |
| ME21 | baseline remission | -0.04 | 0.009 | 0.22 |
| ME12 | baseline remission | 0.03 | 0.02 | 0.22 |
| ME15 | baseline remission | 0.03 | 0.03 | 0.22 |
| ME10 | baseline remission | 0.02 | 0.04 | 0.22 |
| ME22 | Baseline diagnosis | -0.02 | 0.03 | 0.6 |
| *ECT* |  |  |  |  |
| ME3 | time*diagnosis | 0.012 | 0.0002 | 0.004 |
| ME1 | time*diagnosis | -0.003 | 0.0003 | 0.004 |
| ME14 | time*diagnosis | 0.005 | 0.001 | 0.005 |
| ME2 | time*diagnosis | -0.002 | 0.001 | 0.005 |
| ME10 | time*diagnosis | -0.026 | 0.004 | 0.018 |
| ME11 | time*diagnosis | 0.016 | 0.006 | 0.023 |
| ME21 | time*diagnosis | -0.007 | 0.013 | 0.040 |
| ME16 | time*diagnosis | -0.016 | 0.013 | 0.040 |
| ME14 | baseline diagnosis | -0.06 | 0.00003 | 0.001 |
| ME2 | baseline diagnosis | 0.06 | 0.0001 | 0.002 |
| ME23 | baseline diagnosis | 0.05 | 0.001 | 0.010 |
| ME10 | baseline diagnosis | 0.06 | 0.004 | 0.023 |
| ME13 | baseline diagnosis | -0.04 | 0.007 | 0.033 |
| ME3 | baseline diagnosis | -0.06 | 0.008 | 0.033 |
| ME15 | baseline diagnosis | 0.04 | 0.010 | 0.033 |
| ME9 | baseline diagnosis | 0.05 | 0.011 | 0.033 |
| ME7 | baseline diagnosis | -0.04 | 0.013 | 0.033 |
| ME21 | baseline diagnosis | 0.04 | 0.014 | 0.033 |
| ME11 | baseline diagnosis | 0.03 | 0.016 | 0.036 |
| ME1 | baseline diagnosis | -0.04 | 0.018 | 0.036 |
| ME11 | Baseline responder | 0.03 | 0.009 | 0.2 |

## 5. Discussion

We investigated whole blood transcriptome differences as a possible biomarker for rapid-acting antidepressant treatments, including sub-anesthetic ketamine and ECT. Despite the highly significant antidepressant effects observed in these two different TRD treatment cohorts, we failed to detect longitudinal changes in gene expression over time or in association with clinical response after employing rigorous multiple comparison correction methods. Notwithstanding, our results provide some preliminary evidence that cross-sectional pre-treatment transcriptional profiles differ with TRD diagnosis and future treatment response.

### Treatment-Related Changes in Gene Expression

Few prior studies have reported changes in peripheral blood gene expression with antidepressant treatment, primarily focusing on selective serotonin reuptake inhibitors (SSRIs) (39–43). However, these studies show limited overlap in identified genes and have yielded inconsistent findings across cohorts, restricting their generalizability. For example, a large study by Nøhr et al. (2021)(44), which included 281 patients in a placebo-controlled trial of vortioxetine (n=97 placebo), found no significant longitudinal changes in gene expression after 8 weeks of treatment. While some baseline gene expression markers were modestly correlated with treatment response, the overall effects were small. The authors interpreted these results to imply that antidepressant response may exert subtle effects across many genes, such that even relatively large sample sizes may be underpowered to detect these distributed signals. However, SSRIs require several weeks to exert clinical effects, during which time numerous external and internal factors may contribute transcriptional noise. To mitigate the potential confound of these protracted response times, our study instead focused on examining changes in gene function that occur following two rapid-acting antidepressant treatments, including ECT and ketamine, which both produce robust clinical responses within hours to days in individuals with TRD. We expected that these fast-acting treatment modalities would offer a more favorable context for detecting biologically meaningful changes in blood-derived gene expression associated with therapeutic response. Despite observing strong clinical improvements and changes in brain structure(45,46) and function(47–50) previously reported in overlapping samples, we were unable to detect significant or clearly clinically meaningful longitudinal gene expression changes following either intervention.

Though limited prior evidence exists regarding the effects of ECT or ketamine on gene expression, our findings are consistent with those of Ryan et al. (2020)(28), who also reported no significant changes in peripheral blood mRNA following ECT, though they did identify pre-treatment differences between patients and controls. Similarly, Cathomas et al. (2022)(30) reported primarily cross-sectional differences in gene expression, both between patients with TRD (N=26) and unaffected controls (N=21) and between ketamine responders and non-responders.

Our findings together with the limited prior data available suggest that changes in gene expression induced by ketamine and ECT are subtle, and potentially heterogeneous, requiring much larger samples to capture. Another possibility is that our failure to observe significant longitudinal changes in gene expression was due to the sampling of peripheral blood. Although there is some overlap between gene expression in blood and brain—particularly for genes implicated in depression(27)—most transcriptional alterations associated with antidepressant response are expected to occur within brain tissue, which remains inaccessible for *in vivo* experiments(26). That is, while postmortem brain studies have advanced knowledge of brain-specific gene expression, their cross-sectional nature limits their utility in capturing dynamic treatment effects(51). However, emerging models such as brain organoids derived from induced pluripotent stem cells may offer a promising alternative for studying the molecular effects of psychiatric treatments(52). For example, these systems can recapitulate aspects of brain-specific transcription in a controlled setting and enable longitudinal analysis of neural gene expression in response to interventions like ketamine(53). However, organoids remain an imperfect model— they lack the full architectural and functional complexity of the human brain, including vascularization and immune system interactions(54). Moreover, challenges persist in modeling ketamine’s pharmacokinetics, including the ability to cross the blood-brain barrier and drug distribution across different brain regions—factors critical for understanding region-specific transcriptional response(16). These limitations underscore the importance of integrating multiple experimental approaches—spanning peripheral markers, neuroimaging, postmortem studies, and organoid models—to more comprehensively characterize the systemic and dynamic mechanistic effects of rapid-acting antidepressants.

### Depression-related Alterations in Gene Expression

Cross-sectional findings, especially in populations with TRD, may reflect the cumulative burden of prolonged illness, which could be captured in peripheral blood gene expression. Existing studies examining DGE in psychiatric disorders have produced mixed results. For MDD, some studies have identified diagnostic differences in blood-based gene expression(55), while others have reported no significant associations(56). Similarly, findings in bipolar disorder (BD) have been inconsistent. Some studies have reported significant DGE in BD(57,58) but it has been suggested that gene expression patterns observed in BD patients may be primarily due to lithium use(59). While these inconsistencies may be partly attributable to differences in study design and sample size, they are also likely driven by the inherent heterogeneity of mood disorders, including variability in symptom profiles, treatment histories, and underlying neurobiology. These conflicting findings underscore the notion that altered gene expression psychiatric disorders are likely modest and highly context-dependent. It is possible that diagnostic group effects may have been biased by including a treatment-resistant cohort with severe, chronic illness. This may also explain the larger control–ECT differences, as ECT patients had greater symptom severity and comorbidity than the ketamine group (**Supplemental Table 3**). Furthermore, it is possible that we were able to detect some cross-sectional treatment-related effects because individuals who do not respond to ketamine or ECT may have a more unique form of TRD and carry a distinct biological signature; one that is more pronounced, and thus more likely to be reflected in peripheral blood. Alternatively, this profile may reflect the cumulative effects of a prolonged treatment history, with multiple medications potentially leaving a lasting biological imprint. In contrast, responders and remitters may exhibit more transient or subtle molecular changes or perhaps reflect a more plastic or adaptive transcriptional profile. Thus, illness severity, chronicity, and treatment resistance may indeed be reflected in peripheral gene expression; however, these effects require large, well-powered samples to detect reliably.

### Network Analysis Findings

While we observed some significant time-by-diagnosis and time-by-remission interactions after clustering genes into co-expression networks using WGCNA, results were modest (|logFC|<0.05), and the functional classification of genes within significant modules were somewhat uninformative. WGCNA was chosen for its strength in reducing dimensionality, which is important for addressing Type I error and because genes rarely function in isolation. Modules were constructed using data from all time points across all groups (ECT, ketamine, and controls) to capture patterns of gene co-expression across the full sample. Understanding how genes cluster and act together is likely more informative than analyzing individual genes alone. One module, ME21, emerged in several analyses. At baseline, ME21 expression was significantly higher in controls compared to the ECT group (passing FDR correction). In the ketamine group, no results survived FDR correction, but trending results showed non-remitters had higher baseline ME21 expression than remitters. A time-by-remission interaction showed modest ME21 increases in remitters, with no change in non-remitters. Functionally, ME21 was enriched for genes related to immune response and immunoglobulin production, which aligns with hypotheses about immune dysregulation in depression and potential anti-inflammatory effects of ECT and ketamine. However, further inspection revealed that the gene ontology signal was primarily driven by a single gene, IGKV1-9, which was also identified in our differential gene expression analysis. Ideally, biologically meaningful pathway enrichment would be supported by coordinated changes across multiple genes. Given that this is just one gene and the direction of effects was inconsistent, the overall findings, while tempting to interpret within an immune-related framework, are ultimately not robust enough to be considered convincing.

### Limitations

As discussed above, our modest sample size (N=97 participants with TRD, N=35 unaffected controls) may have limited our ability to detect subtle or distributed transcriptional changes, particularly after correction for multiple comparisons. While a subset of controls underwent repeated sampling, a placebo or sham treatment group was not included. Nevertheless, given the absence of robust longitudinal molecular changes—treatment-specific or otherwise—the lack of a placebo is unlikely to have influenced the interpretation of our gene expression findings but is considered a limitation for the specificity of the observed clinical improvements. Notably, the control group was significantly younger than the TRD group, which may have contributed to baseline differences in gene expression and represents a potential confound. Still, the clinical responses we observed are consistent with the well-established efficacy of ketamine and ECT(12,13,60) supporting the interpretation that these changes were likely attributable to active treatment.

Regarding timing of blood collection, Blood was collected 24 hours post-treatment, when antidepressant effects are typically strongest for ketamine and ECT. However, some rapid and transient transcriptional changes may have occurred within hours of treatment and were not captured. That said, if ketamine or ECT exert meaningful downstream effects on immune or inflammatory pathways—as suggested by prior work—one might expect such signals to persist for at least 24 hours and thus remain detectable. The absence of robust effects 24 hours post-treatment raises further questions about the magnitude or consistency of these peripheral molecular signatures.

### Conclusion

Our findings may benefit future studies investigating the molecular mechanisms of antidepressant response. The lack of robust longitudinal changes in peripheral gene expression suggests that RNA-seq may have limited immediate translational utility, and alternative approaches—such as DNA methylation, proteomics, or multimodal integration—may prove more informative. While ketamine and ECT produced clear clinical benefits in TRD, we failed to detect clear and consistent transcriptional signatures in peripheral blood relating to treatment. Modest cross-sectional differences hint at underlying biological distinctions, but larger discovery studies with greater statistical power and refined phenotyping will be critical for identifying reliable biomarkers.

## Data Availability

Genetic data for this study can be found in the Gene Expression Omnibus database repository.

## 6. Acknowledgements

Research reported in this research was supported by the NIMH Grants No. **U01MH110008** (KLN, RTE, EC) and **R01 MH115676** (RAO), UCLA Neurobehavioral Genetics NINDS **T32NS048004** (AZP), the UCLA Intercampus Medical Genetics training program USHHS Ruth L. Kirschstein Institutional NRSA # **T32GM008243** (AZP).

