## Supplemental Files for "Transcriptional profiling of antidepressant ketamine and electroconvulsive therapy treatment"

### Demographics Tables

**Supplemental Table 1**. Mean age and sex distributions of participants with available plasma samples at each time point during ECT or ketamine treatment

| | N | Age | Percent Female |
|---|---|---|---|
| *Ketamine* | | | |
| Baseline | 60 | 39.5±11.2 | 46.7% |
| 24 hrs post 1st infusion | 39 | 38.7±10.9 | 35.9% |
| 24 hrs post 4th infusion | 52 | 39.3±10.7 | 48.1% |
| 5-week follow up | 37 | 40.1±10.2 | 43.2% |
| *ECT* | | | |
| Baseline | 37 | 41.6±16.0 | 67.6% |
| <1 week after the ECT index | 32 | 40±16.0 | 62.5% |
| 3-month follow up | 20 | 37.7±13.3 | 60% |

**Supplemental Table 2.** Race and education of study participants

| Education (%) | | | | Race (%) | | | |
|---|---|---|---|---|---|---|---|
| | KET | ECT | HC | | KET | ECT | HC |
| 10th grade (2) | 1.54% | 0% | 0% | White, Non-hispanic (1) | 70.77% | 73.17% | 25.64% |
| High School Graduate (3) | 3.08% | 2.44% | 2.56% | White, Hispanic/latino (2) | 7.69% | 2.44% | 17.95% |
| Some College (5) | 27.69% | 26.83% | 20.51% | Asian (3) | 10.77% | 7.32% | 17.95% |
| Associates Degree (4) | 4.61% | 12.19% | 2.56% | Black (4) | 1.54% | 4.88% | 17.95% |
| Bachelor's Degree (6) | 33.85% | 29.27% | 46.15% | More than one race (5) | 3.08% | 4.88% | 7.69% |
| Masters Degree (7) | 20% | 14.63% | 17.95% | Other (6) | 4.61% | 7.32% | 12.82% |
| Professional Degree (9) | 6.15% | 14.63% | 7.69% | Not reported (7) | 1.54% | 0% | 0% |
| Doctoral Degree (8) | 3.08% | 0% | 2.56% | | | | |

**Supplemental Table 3.** Concurrent medication and psychiatric comorbidities. All participants were diagnosed with treatment resistant depression and were experiencing a major depressive episode at the time of the study. Other diagnoses were defined using a structured clinical interview for DSM (SCID).

| | **Subcategory** | **KET (%)** | **ECT (%)** |
|---|---|---|---|
| **Medications** | SSRI | 30.77 | 24.39 |
| | SNRI | 30.77 | 26.82 |
| | Other Antidepressants | 52.31 | 43.90 |
| | Lithium | 1.53 | 12.19 |
| | Benzodiazepines* | 29.23 | 31.70 |
| | Anticonvulsants | 24.61 | 39.02 |
| | Atypical Antipsychotics | 20.00 | 36.58 |
| | Stimulants | 24.61 | 4.88 |
| | No Current Psychotropic Medication | 20.00 | 0.00 |
| **Other Diagnoses** | MDD with Bipolar Features | 3.39 | 24.39 |
| | PTSD | 22.03 | 46.34 |
| | Anxiety | 62.71 | 65.85 |
| | History of Cannabis Dependence | 10.17 | 14.63 |
| | History of Alcohol Dependence | 27.12 | 41.46 |
| | History of Nicotine Dependence | 9.23 | 17.07 |

*Patients were requested to withhold benzodiazepines >24 hrs before treatment sessions and blood draws

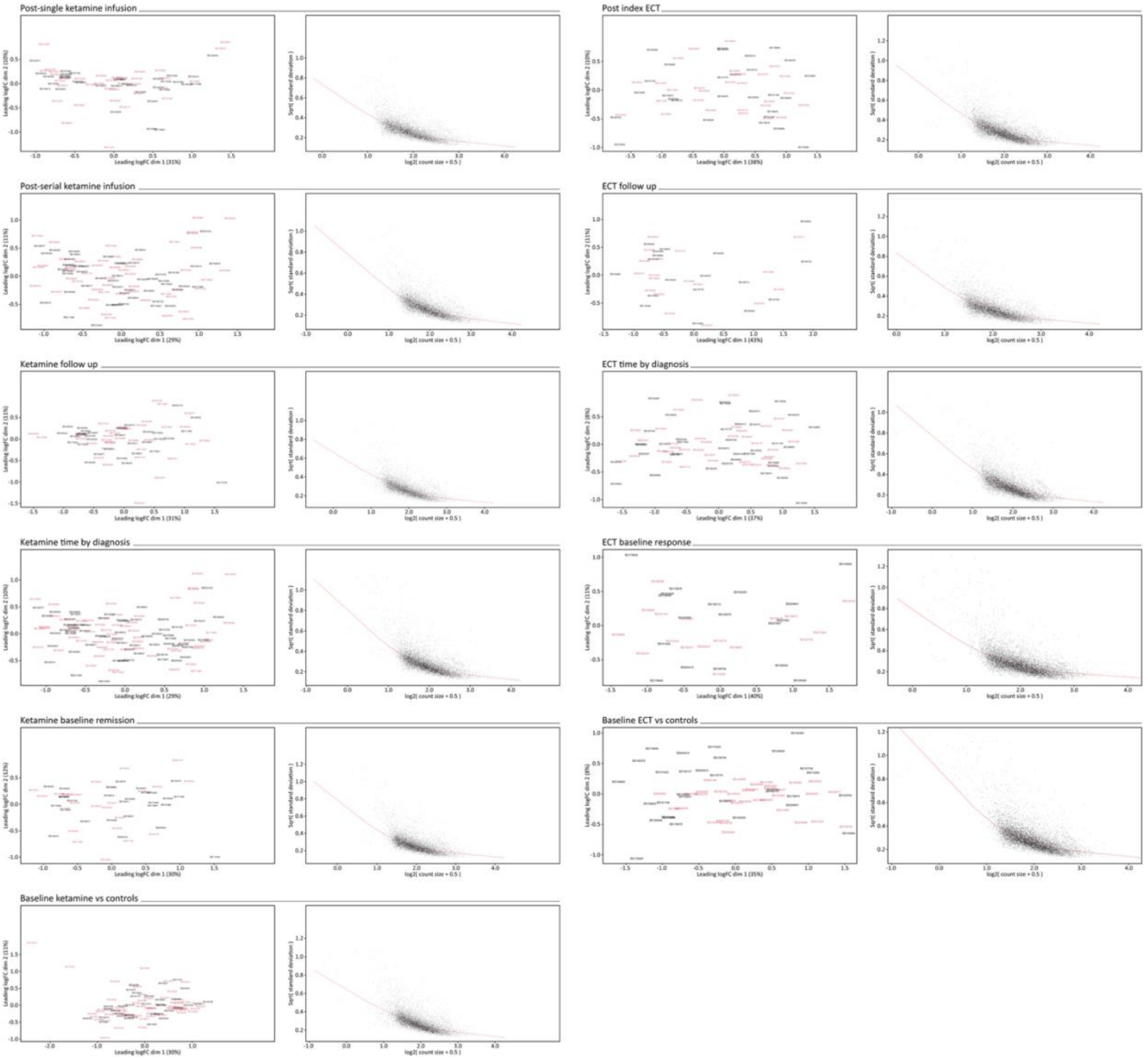


**Supplemental Figure 1.** Multidimensional scaling (MDS) plot and limma-voom mean-variance trend J-plots for each analysis.

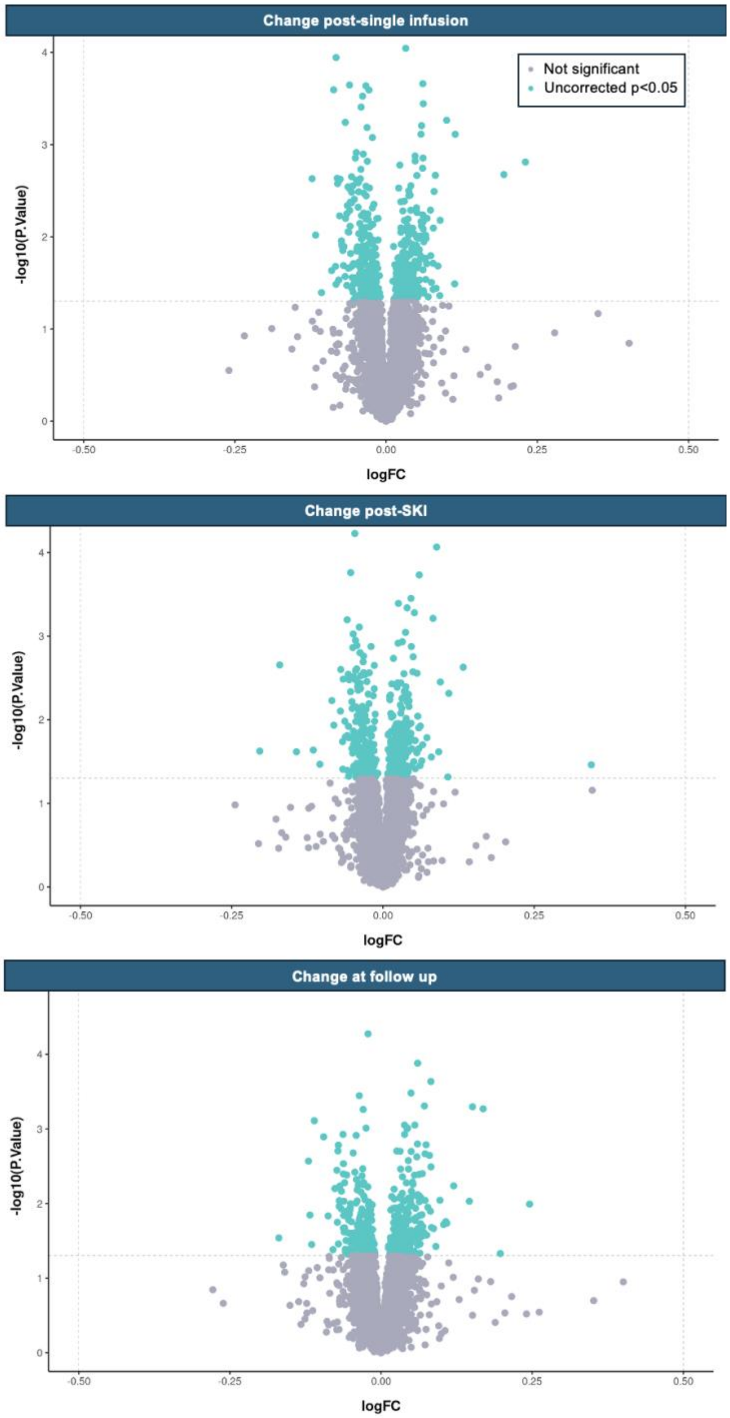


**Supplemental Figure 2.** Volcano plots for pairwise DGE analysis in the ketamine sample. Comparisons are base with baseline and each timepoint.

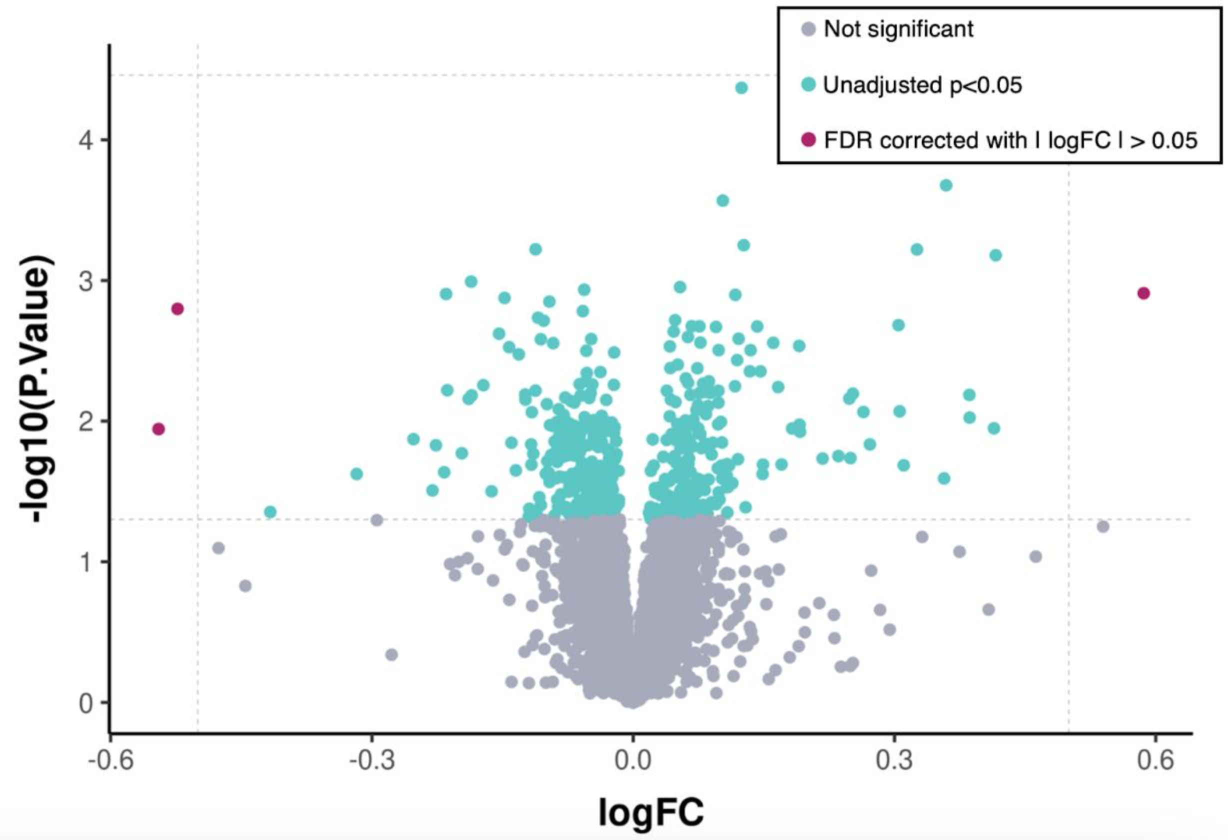


**Supplemental Figure 3.1**. Ketamine sample volcano plot for results comparing time*remission status when comparing from baseline to SKI.

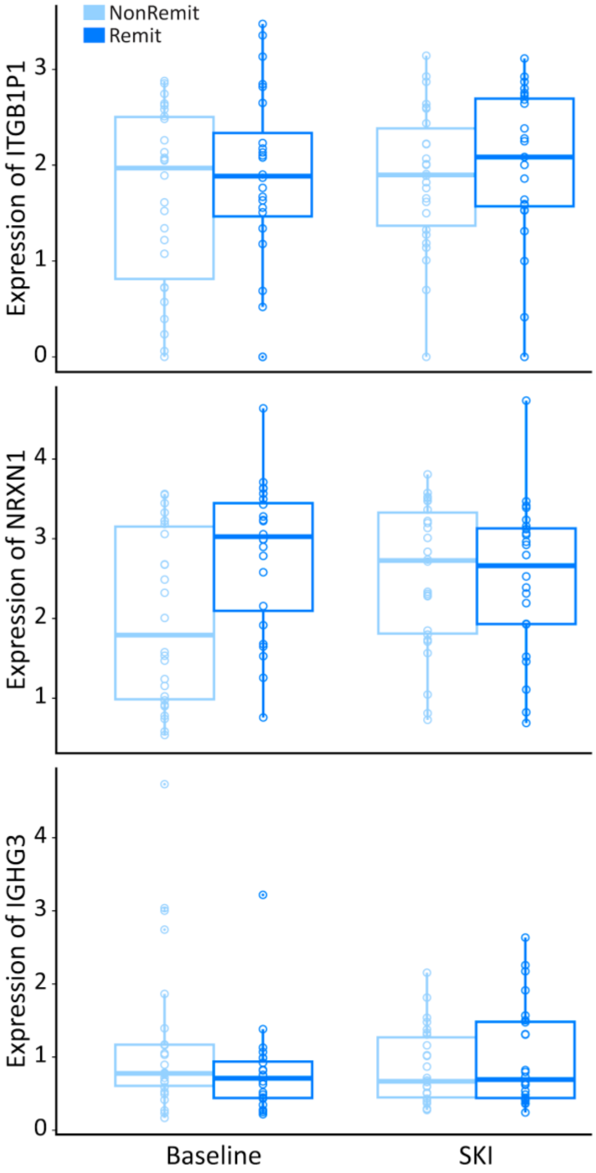


**Supplemental Figure 3.2**. Boxplots showing differences between ketamine patients defined as remitters and non-remitters at baseline and following serial ketamine for trending genes extracted from the time*remission interaction.

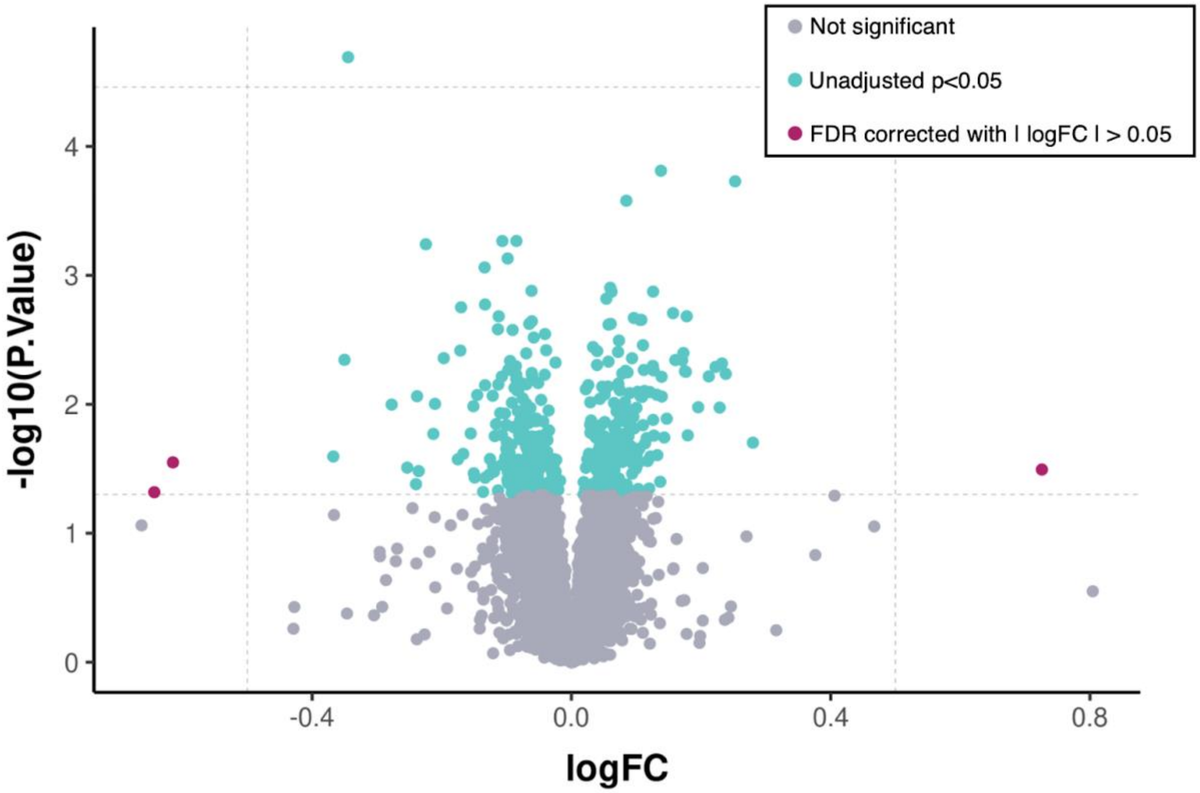


**Supplemental Figure 4.1.** Ketamine sample volcano plot for results comparing time*diagnosis.

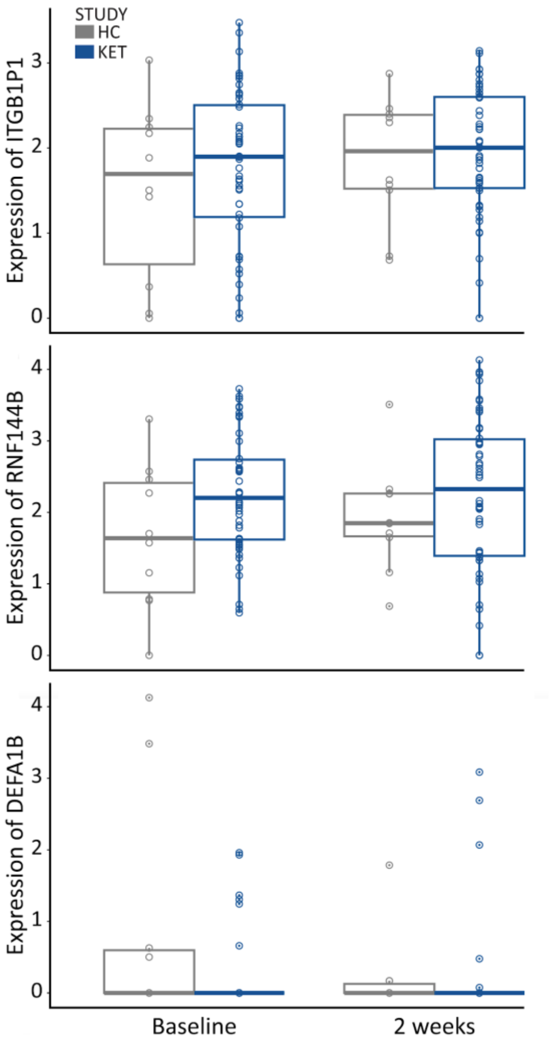


**Supplemental Figure 4.2.** Boxplots showing differences between ketamine patients and controls at baseline and after 2-weeks for controls and serial ketamine for patients for genes surviving FDR correction from the significant genes for time*diagnosis analysis.

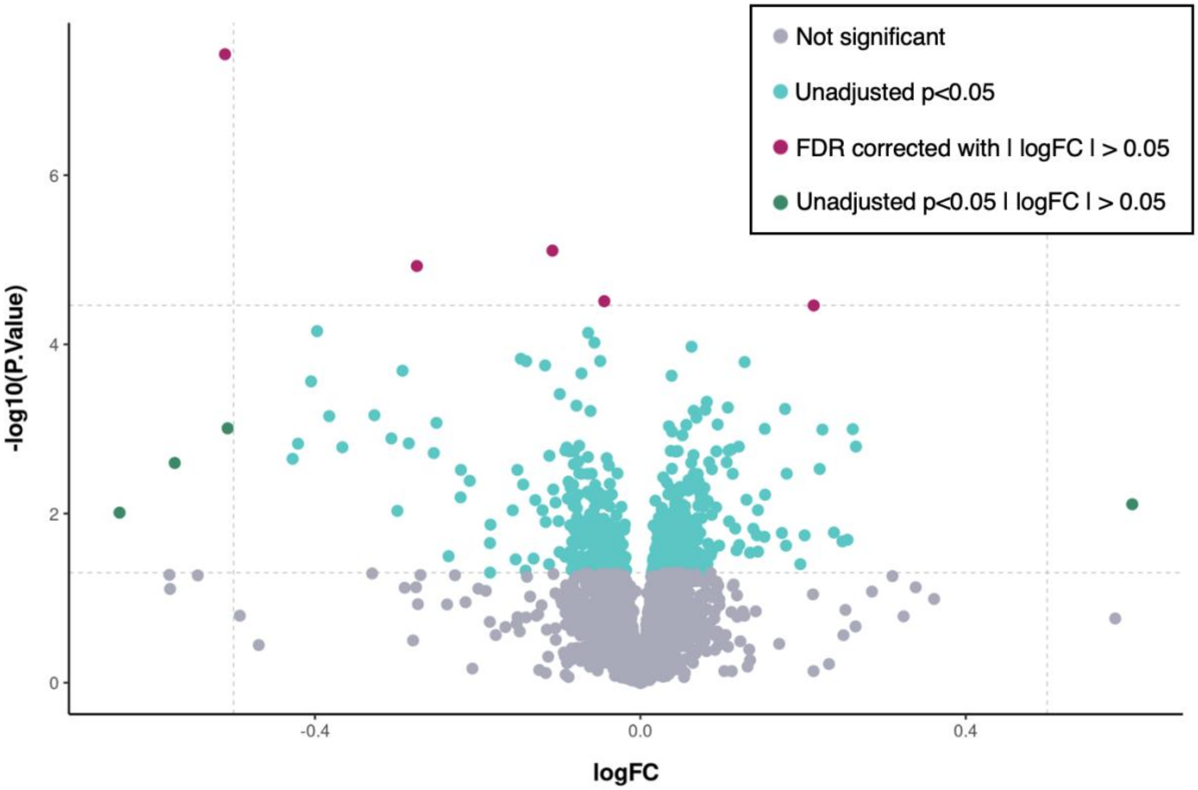


**Supplemental Figure 5**. Ketamine sample volcano plot for results comparing remitters versus not remitters at baseline.

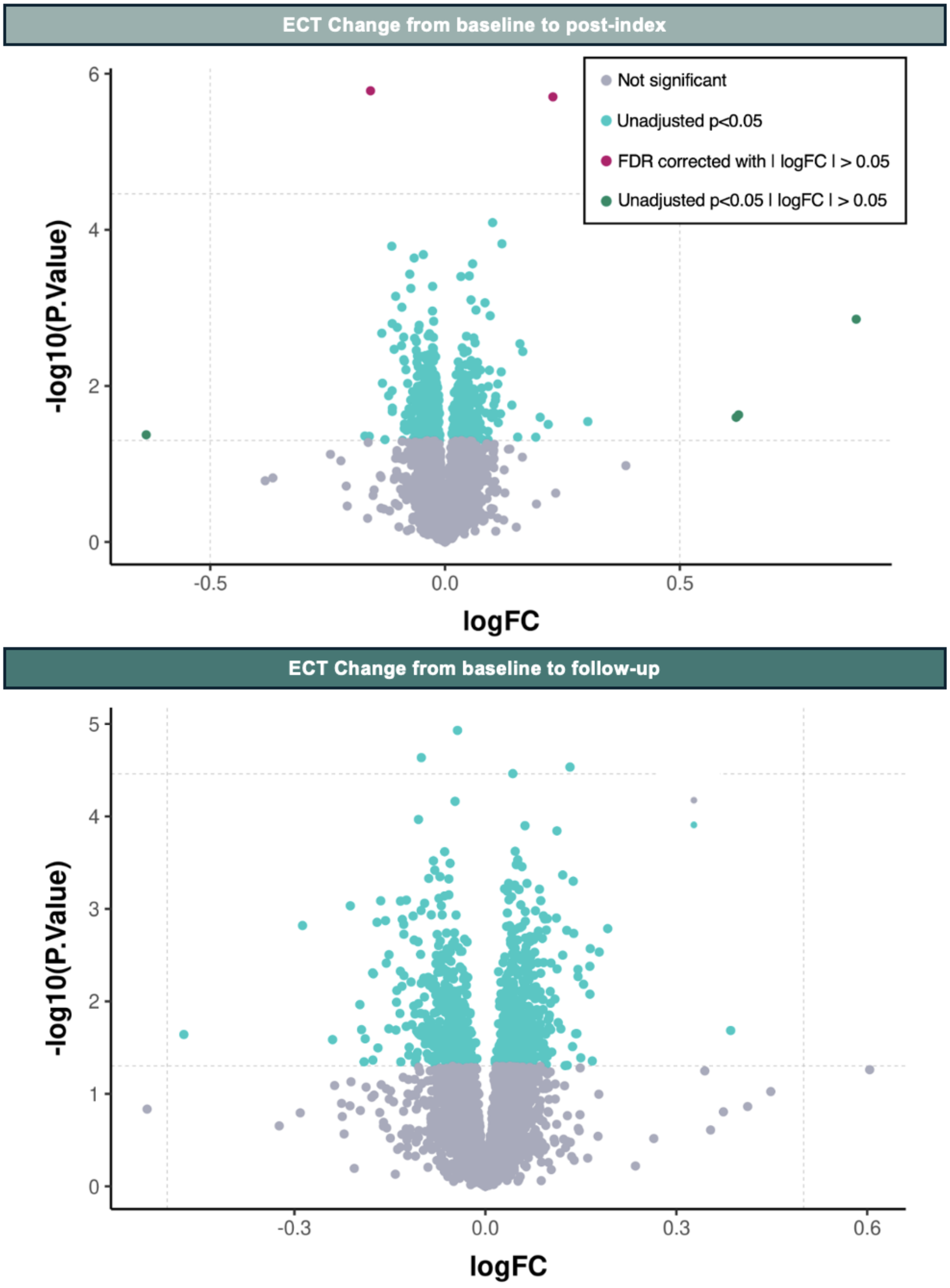


**Supplemental Figure 6**. ECT sample volcano plot for results comparing longitudinal changes from baseline to post-index.

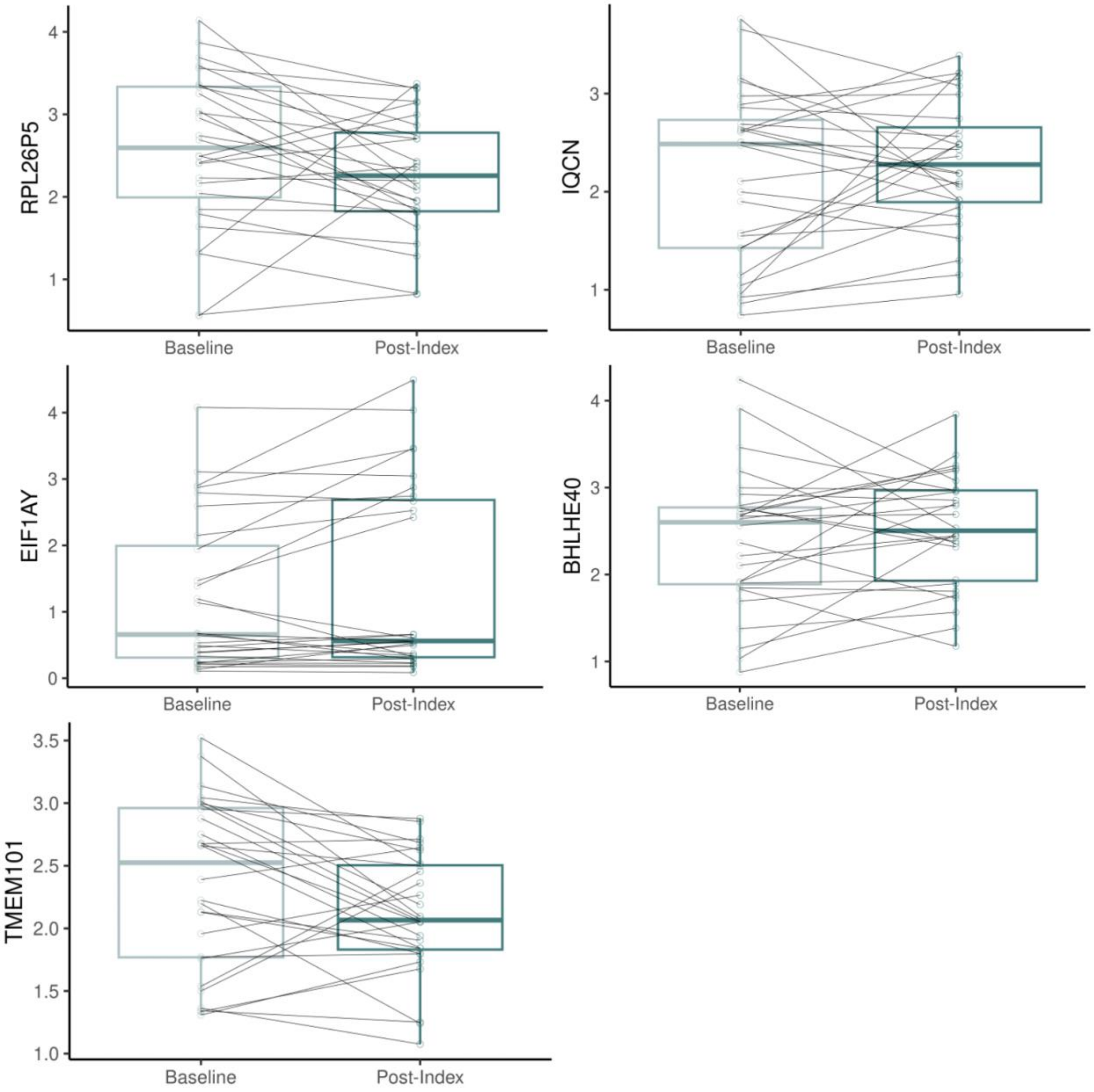


**Supplemental Figure 7**. Boxplots showing genes with trending changes in the ECT sample from baseline to post-index.

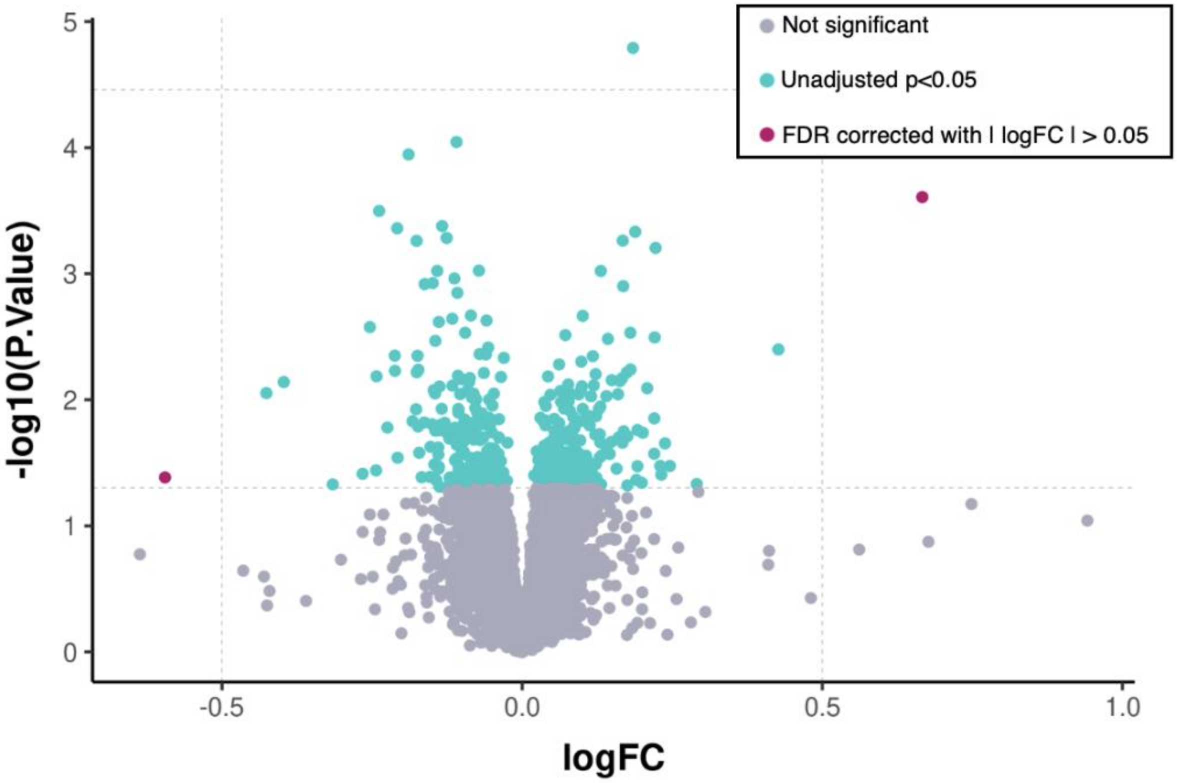


**Supplemental Figure 8**. ECT sample volcano plot for results investigating time*diagnosis.

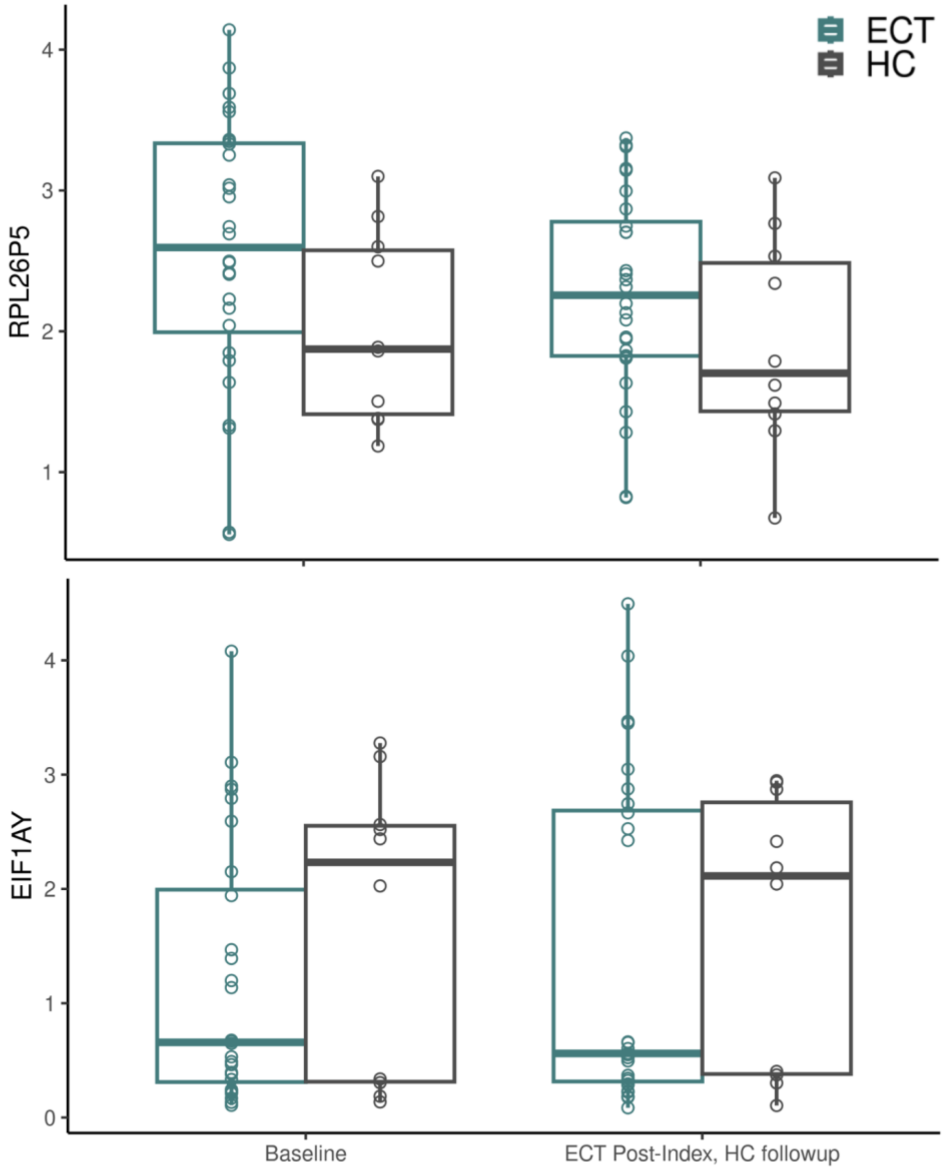

**Supplemental Figure 9.** Boxplots showing longitudinal changes for ECT (post-index) and controls (follow up) for genes that were trending in significance for a time*diagnosis interaction.

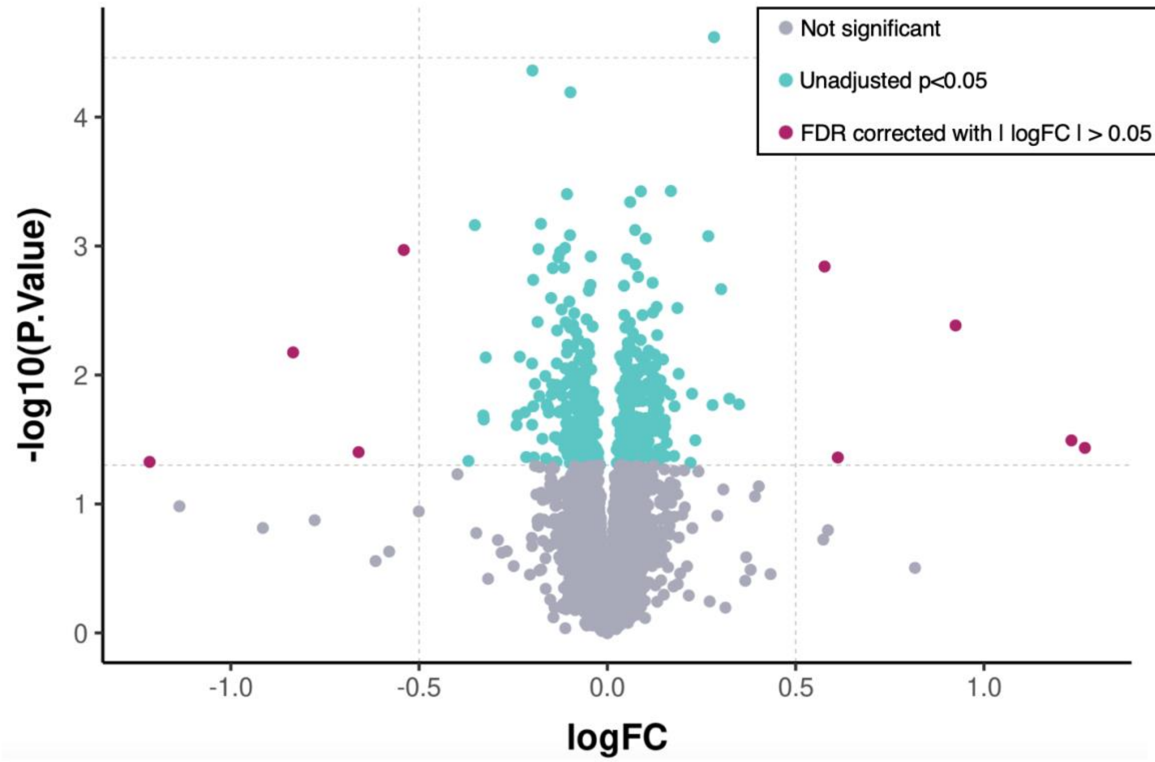


**Supplemental Figure 10.** ECT sample volcano plot for results comparing responders and non-responders at baseline.

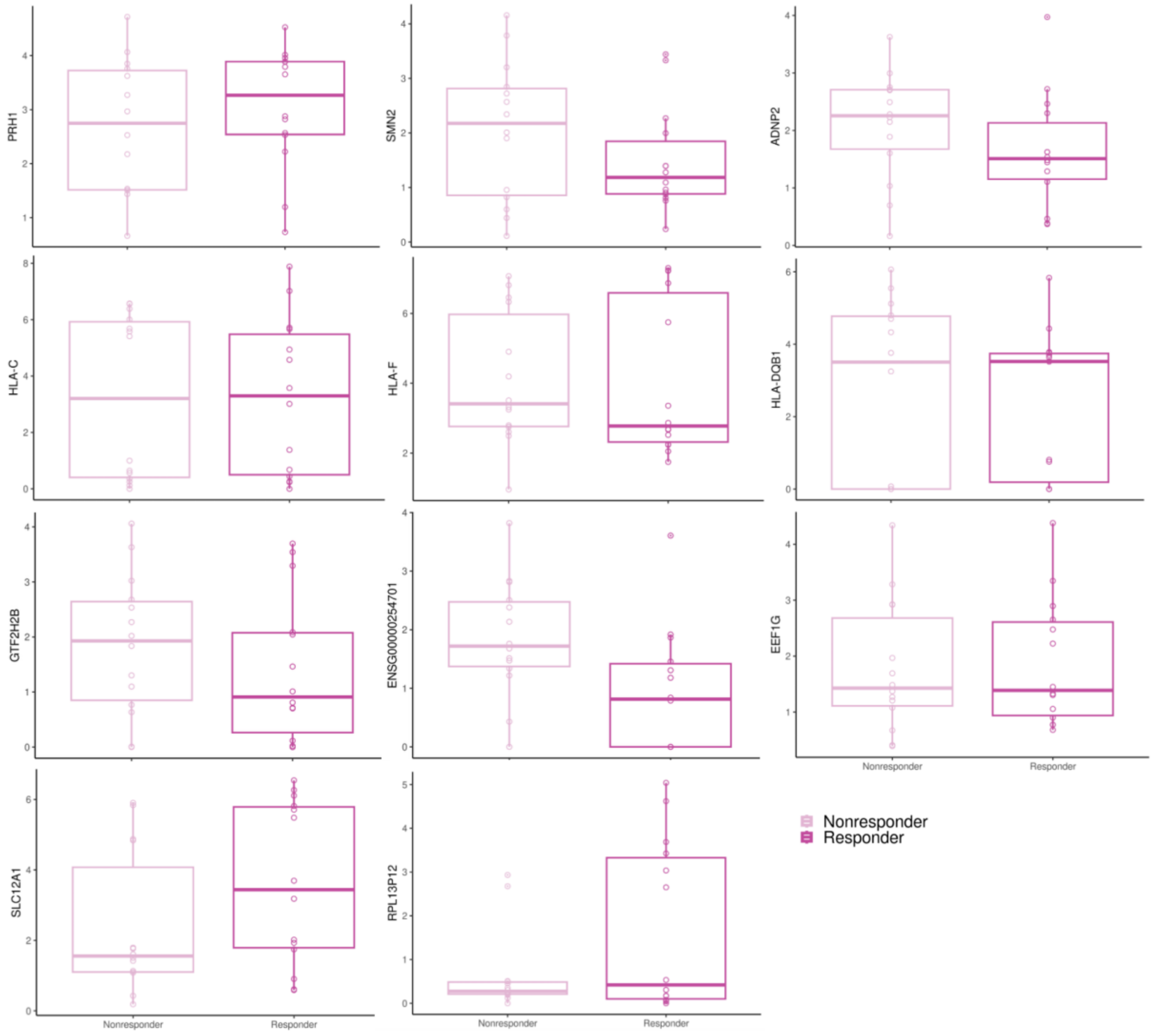


**Supplemental Figure 11.** Boxplots of trending genes differentially expressed when comparing ECT responders and non-responders at baseline.

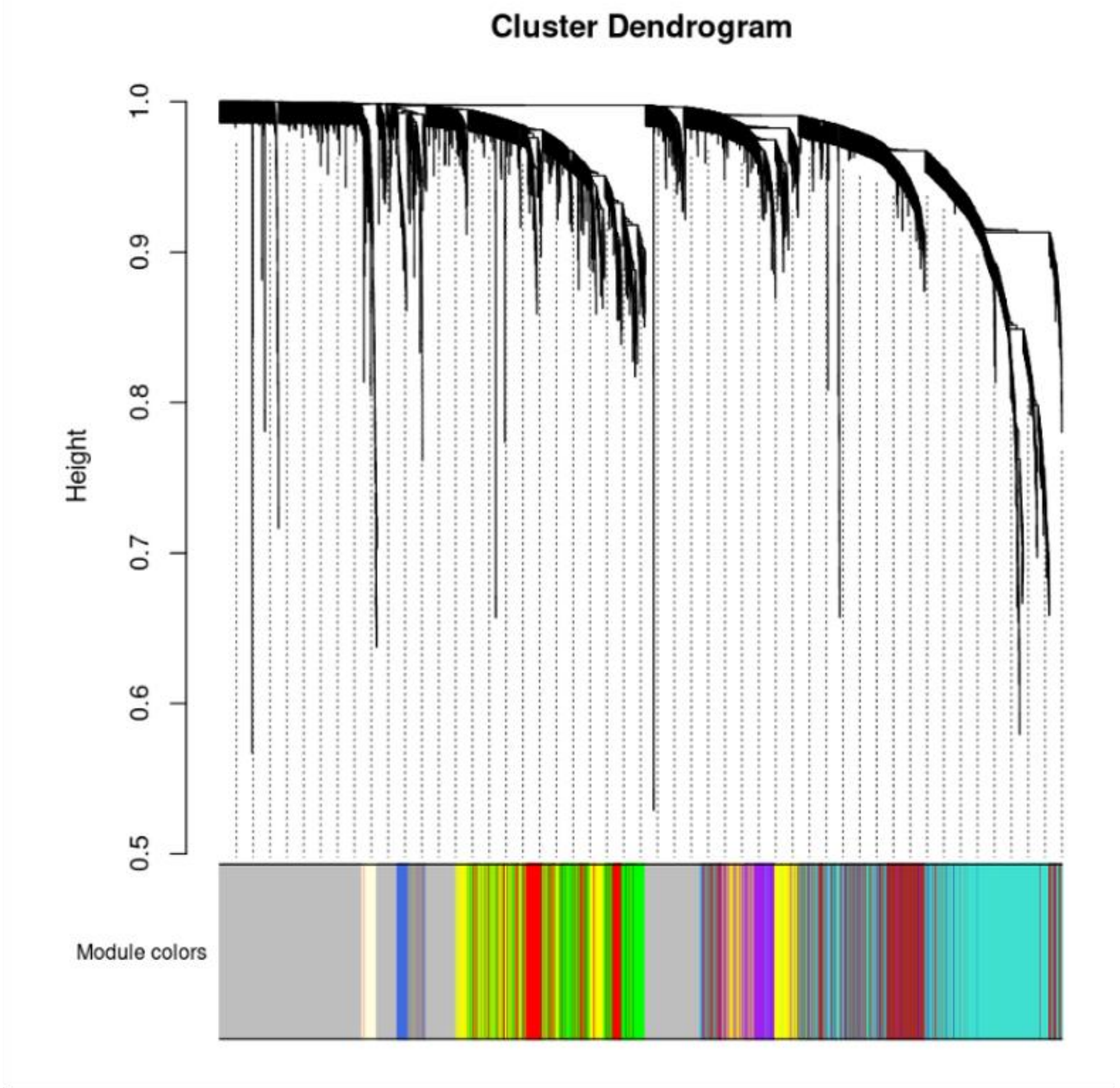


**Supplementary Figure 12**. Cluster dendrogram for WGCNA created using data across time points and groups (ECT, Ketamine, controls).

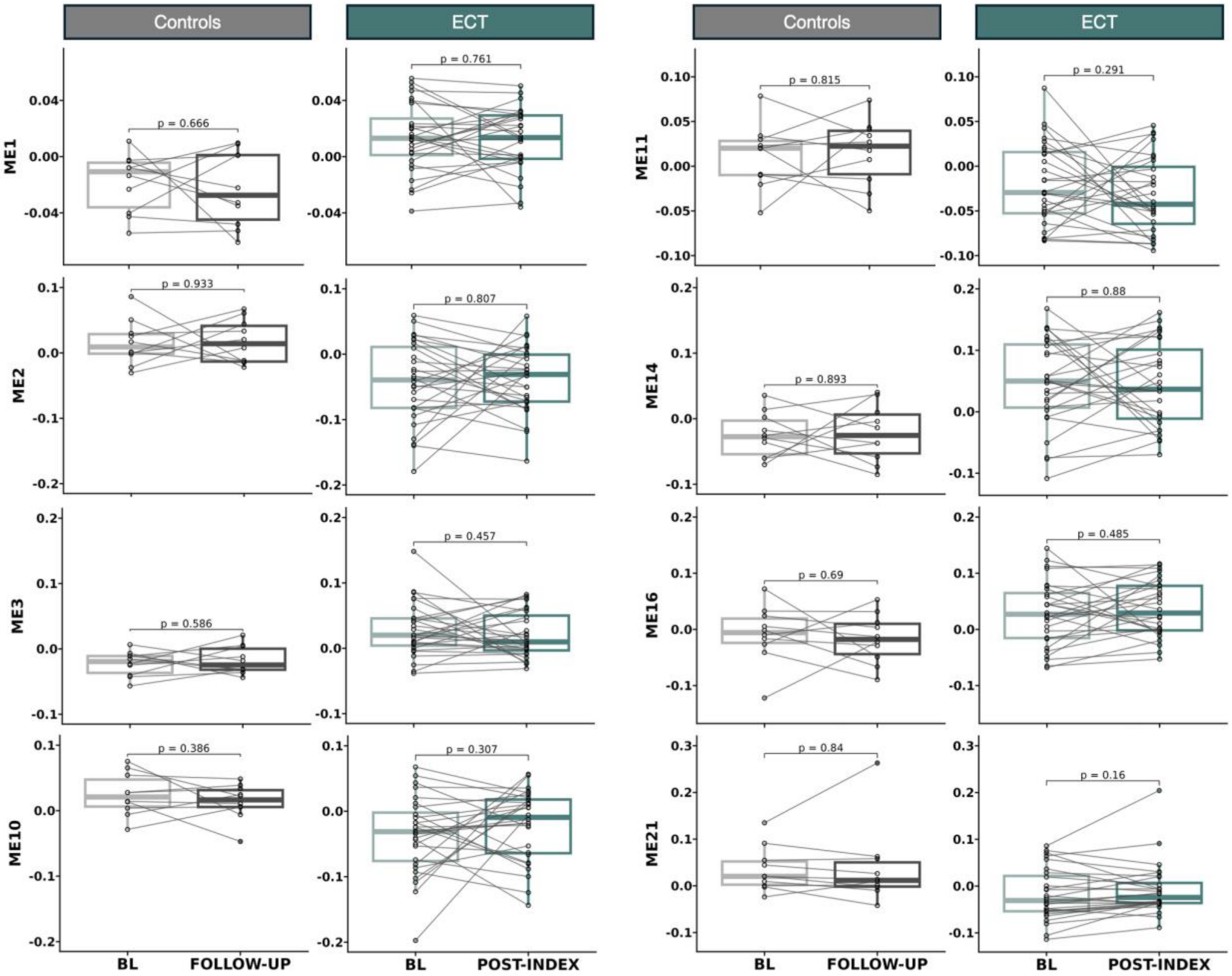


**Supplemental Figure 13.** Boxplots for significant eigengene findings for results from the time*diagnosis analyses for ECT.

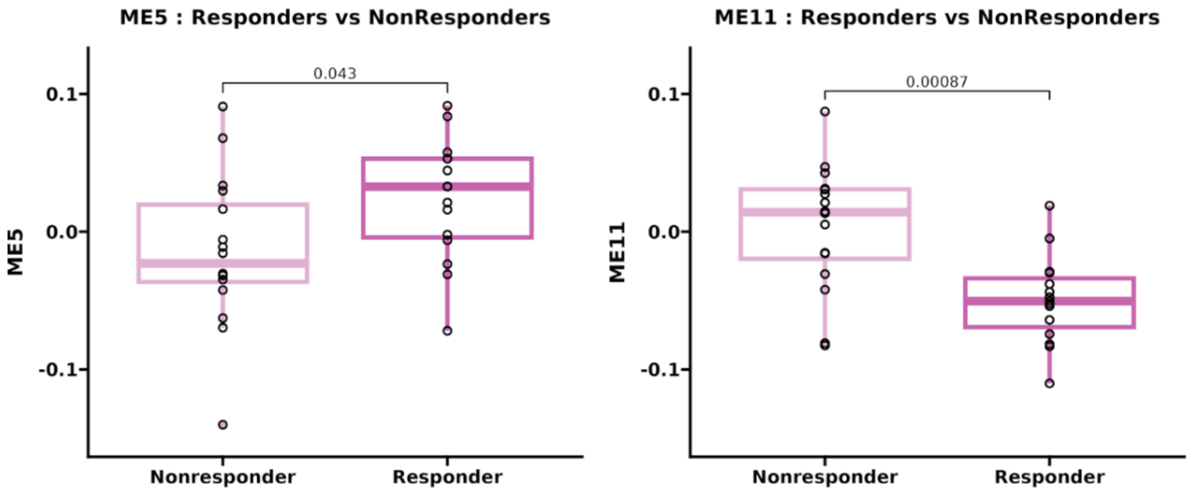


**Supplemental Figure 14.** Boxplots for significant baseline comparisons of responders to ECT.

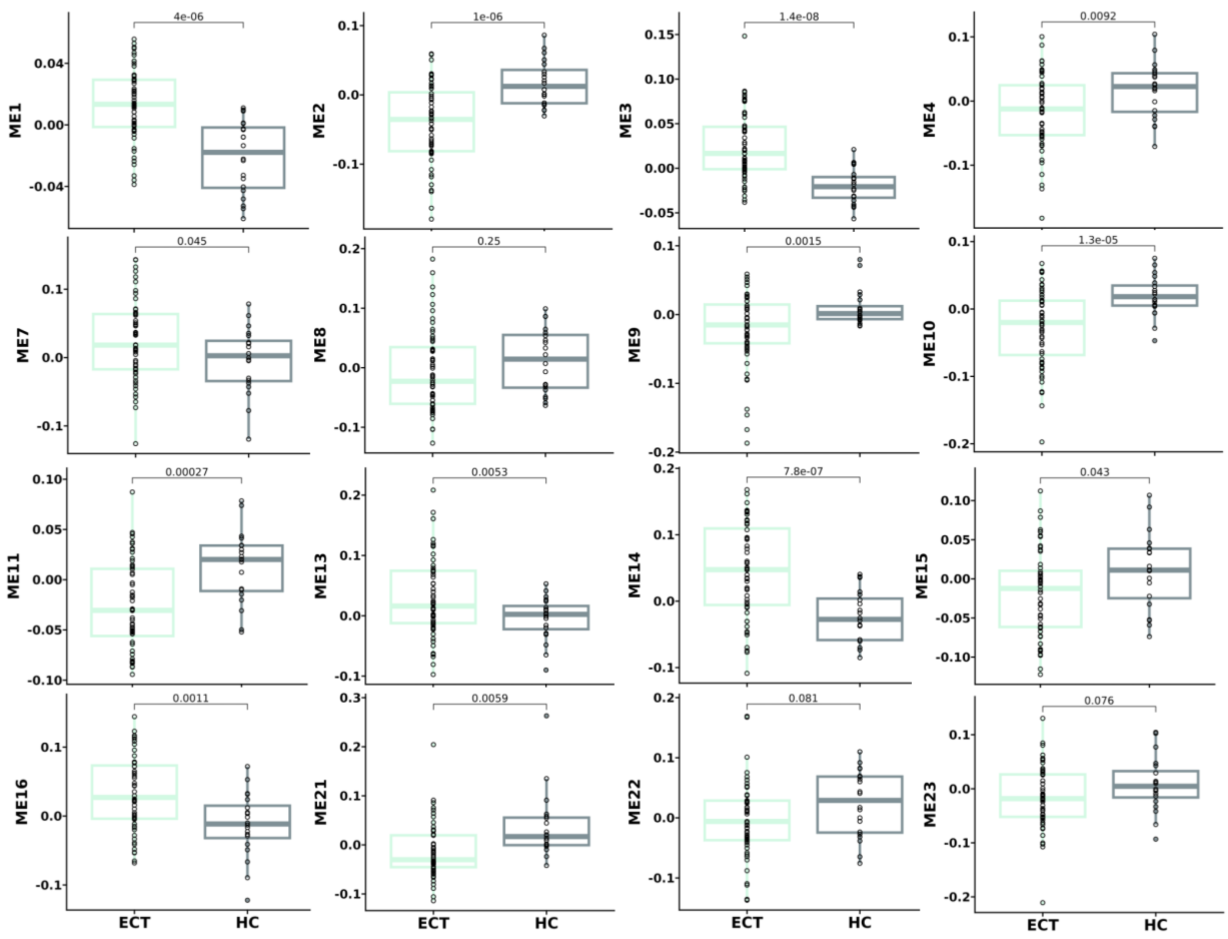


**Supplemental Figure 15.** Boxplots for significant eigengene differences between ECT patients and HC at baseline.
